# 12-weeks of Slow Breathing Exercises Reduces Blood Pressure among Healthy Normotensive Subjects

**DOI:** 10.1101/2022.08.30.22279389

**Authors:** Alfredo Gamboa, Hui Nian, Emily C. Smith, Sachin Paranjape, Katrina Nelson, Robert Abraham, Andre Diedrich, Gurjeet Birdee

## Abstract

Slow breathing exercises are a key component to many mind-body practices used for stress reduction and modulation of autonomic tone, and it has been shown to reduce blood pressure acutely. Long-term effects on blood pressure and autonomic tone are not well documented. We studied 95 healthy participants (41±4 years, 76% female) who performed slow breathing exercises for 12 weeks and examined the effect of slow breathing on systolic blood pressure, diastolic blood pressure, and autonomic tone.

At baseline average BP was 105±11/67±8 mmHg. Among the 11 participants with elevated blood pressure, BP was 126±11.0/ 80±5 mmHg. Our comparison group, that received no intervention, consisted of 30 participants with baseline mean BP 125±19/ 71±10 mmHg.

SBP and DBP decreased significantly (−2.4±7.3 and -1.6±5.5 mmHg, respectively) at 12 weeks for all participants who practiced slow breathing. The comparison group showed no significant changes in blood pressure. Blood pressure reduced further among slow breathing participants with baseline SBP over 120mmHg and/or DBP over 90mmHg (−10.3±7.9 and -3.8±5.5 mmHg, respectively). In our regression model, baseline SBP was associated with further decreases in SBP from baseline to 12 weeks. No significant changes were observed in spectral analyses from baseline to 12 weeks or correlations between spectral analyses in blood pressure.

In conclusion, 12-weeks of slow breathing exercises resulted in a significant reduction of blood pressure in the absence of significant changes in autonomic tone as measured by spectral analyses. Further research into the mechanisms and effectiveness of slow breathing on cardiovascular health is needed.

## Introduction

Almost half the population in the United States has hypertension with only one quarter having their blood pressure under control ^1^. Half of hypertensive patients are either not taking or not prescribed antihypertensive medications. For some patients, pharmacologic treatments are limited by side effects, adherence, and/or efficacy. Lifestyle treatment options for hypertension such as diet and physical activity modifications are recommended but also limited by patient adherence. Additional non-pharmacologic treatments for hypertension may augment current therapeutic options and help reduce the burden of this condition.

Slow breathing exercises are used for health by 12% of adults in the United States ^2^. In research, slow breathing has been defined as a respiratory rate less than 10 breaths a minute. Research suggests that slow breathing decreases sympathetic and increases parasympathetic tone. This may be partially mediated through alteration of intra-thoracic pressures ^3–5^, stimulation of arterial and cardiopulmonary baroreceptors ^6,7^ and afferent pulmonary stretch receptors^3,4,6,7^, or through central interactions between respiratory and cardiovascular centers in the brainstem modulating gating of vagal activity during breathing^8^. Yoga, a popular form of exercise in the United States, includes an extensive and detailed approach to slow breathing techniques for health called *pranayama*. Yoga’s slow breathing techniques acutely lower blood pressure, which may be mediated through modulation of the autonomic nervous system. Yoga slow breathing requires self-direction to slow breathing. An alternative is device-guided breathing, such as RESPeRate (InterCure, Ltd. London, UK) which through biofeedback has users slow breathing. The Food and Drug Administration has approved RESPeRate for stress and blood pressure reduction. Though high-quality studies that measure the effects of yoga or device-based slow breathing on blood pressure are lacking, the Food and Drug Administration has approved RESPeRate for stress and blood pressure reduction ^9^ even though there seems to be insufficient evidence to warrant its widespread use ^10^.

There is agreement that most forms of essential hypertension are characterized by increased sympathetic and decreased parasympathetic tones ^11^. This is especially true in early-stage hypertension before secondary end-organ damage ^12^. We have previously conducted a clinical trial among healthy adults (n=99) who performed slow breathing exercises for 12 weeks to examine effects on psychological and physiological stress. We observed significant changes in psychological stress as measured by PROMIS-Anxiety, but non-significant changes physiological stress measured as heart rate variability. The objective of the present study was to assess the effects of slow breathing on blood pressure. We hypothesized that slow breathing for 12 weeks would have a significant effect on reducing both systolic and diastolic blood pressure. We also examined if any changes in blood pressure correlate with changes in autonomic tone. In addition, we explored if two slow breathing techniques (length of inhale=exhale, I=E, versus length of inhale<exhale, I<E) produce differential effects on blood pressure.

## Methods

This study was approved by the Vanderbilt University Institutional Review Board. Written consent was obtained from each participant and the study was registered in ClinicalTrials.gov prior to enrollment (NCT02870868). The general study design and protocol was identical to what we previously described. Briefly, we enrolled healthy adults aged 30 to 60 years without a diagnosis of hypertension. Patients were excluded if on anti-hypertensive medications. Briefly (figure 1) there was an initial two-weeks screening period to determine eligibility followed by randomization I=E or I>E slow breathing for 10 weeks. During the screening phase, participants attended 2 private classes with a yoga teacher to learn basic slow breathing. Participants were asked to practice daily. Participants were included in the second phase if after the first two weeks: 1) they practiced 3 times or more a week, and 2) had a breath rate between 3-8 breaths per minute. The second phase consisted of 10 -weeks of continued slow breathing training with week-to-week progressions based on a pre-specified 10-week protocol. The slow breathing interventions were delivered individually in a private room at VUMC. Participants were asked to practice slow breathing at home in addition to weekly classes. All eligible subjects were seen at Vanderbilt’s Autonomic Dysfunction Center where standard blood pressure and autonomic testing for spectral analysis of heart rate and blood pressure variability was performed in the morning at baseline before randomization and 12 weeks.

**Figure 1.**
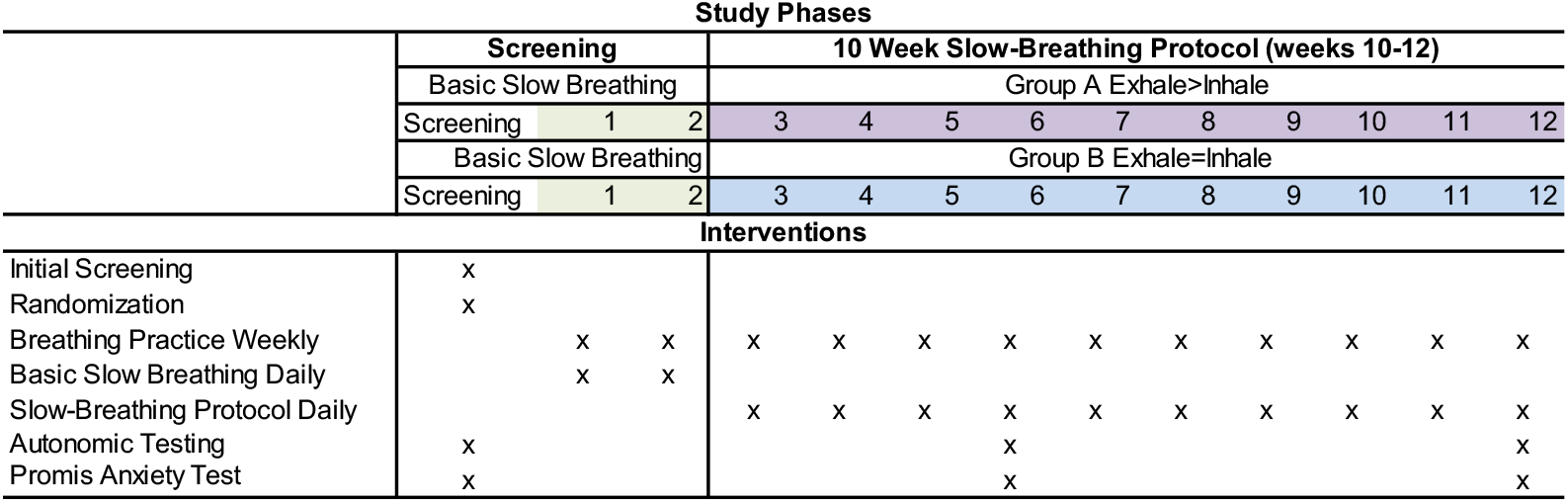
Description of the Protocol used for the Intervention Group.

We defined elevated blood pressure at baseline based on the standard definition of Stage 1 Hypertension being a systolic blood pressure higher than 120 mm Hg or diastolic blood pressure greater than 80 mm Hg. For control, we used data derived from another of our studies that measured blood pressure in normotensives (n=14) and hypertensive (n=15) measured on two separate visits at least one month apart. Hypertensive subjects were included if they were on antihypertensive medication or had at least three documented blood pressure measurements >140/90 mm Hg. For the study visits, antihypertensive medications were withdrawn for at least seven days or five half-lives. Both control groups were asked to abstain from drinking caffeinated drinks at least two days prior to testing. These control groups did not have standard autonomic testing done in the same manner but were included for comparison to rule out an effect of acclimatization to the laboratory environment by the slow breathing participants.

Assessments were performed at the Vanderbilt Autonomic Dysfunction Center as per standardized protocol at baseline and 12 weeks. Participants fasted 8 hours before testing. Medications or supplements that interfere with autonomic tone were held prior to assessments. Subjects were studied in the morning after at least 30 minutes of quiet rest in the supine position, to allow for familiarization with the study procedures and the laboratory environment. Autonomic function was assessed by standardized autonomic testing as previously described ^13,14^. This included a standing stress test, respiratory sinus arrhythmia, and Valsalva maneuver. Sinus arrhythmia was measured during controlled breathing (at 6 breaths/min) and was obtained by dividing the longest to the shortest RR intervals. Brachial blood pressure and heart rate were determined at intervals, using an automated cuff-oscillometric sphygmomanometer (VitalGuard 450C, Ivy Biomedical, Branford, CT), and continuously with the finger clamp method (Nexfin, BMEYE, Amsterdam, the Netherlands) and ECG (VitalGuard450C). Cardiovascular signals were digitized using a Windaq system (DA-220; DATAQ Instruments). Spectral analysis was performed following Task Force recommendations ^15^ and analyzed as previously described^16^.

## Statistical Methods

Descriptive statistics of participant characteristics are presented as median, quartiles, and mean ± SD for continuous variables and frequency for categorical variables. Between-group comparisons were conducted using Wilcoxon rank sum test for continuous variables and Pearson’s chi-squared test for categorical variables. Within-subject change in blood pressure from baseline to 12 weeks was tested using Wilcoxon signed-rank test. To investigate the effect of baseline blood pressure on blood pressure change at 12 weeks of intervention, separate linear regression models were fitted for SBP, DBP and MAP. Blood pressure change from baseline to 12 weeks was used as the response variables; baseline blood pressure, intervention groups as well as interaction between baseline BP and intervention groups were included as independent variables. Correlations between BP change and 12-week autonomic measures / change in autonomic measures were evaluated using Spearman’s correlation coefficient.

## Results

Our slow breathing study enrolled 99 healthy adults of whom 95 participants had complete blood pressure and autonomic measurements at baseline and 12 weeks. Our comparison group consisted of 30 participants who had measurements at baseline and about four weeks. Table 1 shows descriptive demographics on screening of all subjects enrolled. For those in the slow breathing study, the mean SBP was 105 ± 11 and DBP of 67 ± 8. Among the 11 participants with elevated blood pressure, the mean SBP was 126 ± 11.0 and DBP 80 ± 5. Our comparison group, that received no intervention, consisted of 30 participants with mean SBP of 125 ± 19 and mean DBP 71 ± 10. Fourteen of these participants had hypertension with a mean SBP of 141 ± 13 and DBP 76 ± 8. By design there are significant differences in blood pressure among groups. We also found that participants with higher blood pressure were older and had higher body mass indexes. In addition, the intervention group subjects with high blood pressure had significantly higher resting heart rate (70±9 vs. 64±9 bpm for the elevated BP group versus the Normal BP group). No other demographic differences were found.

**Table 1.**
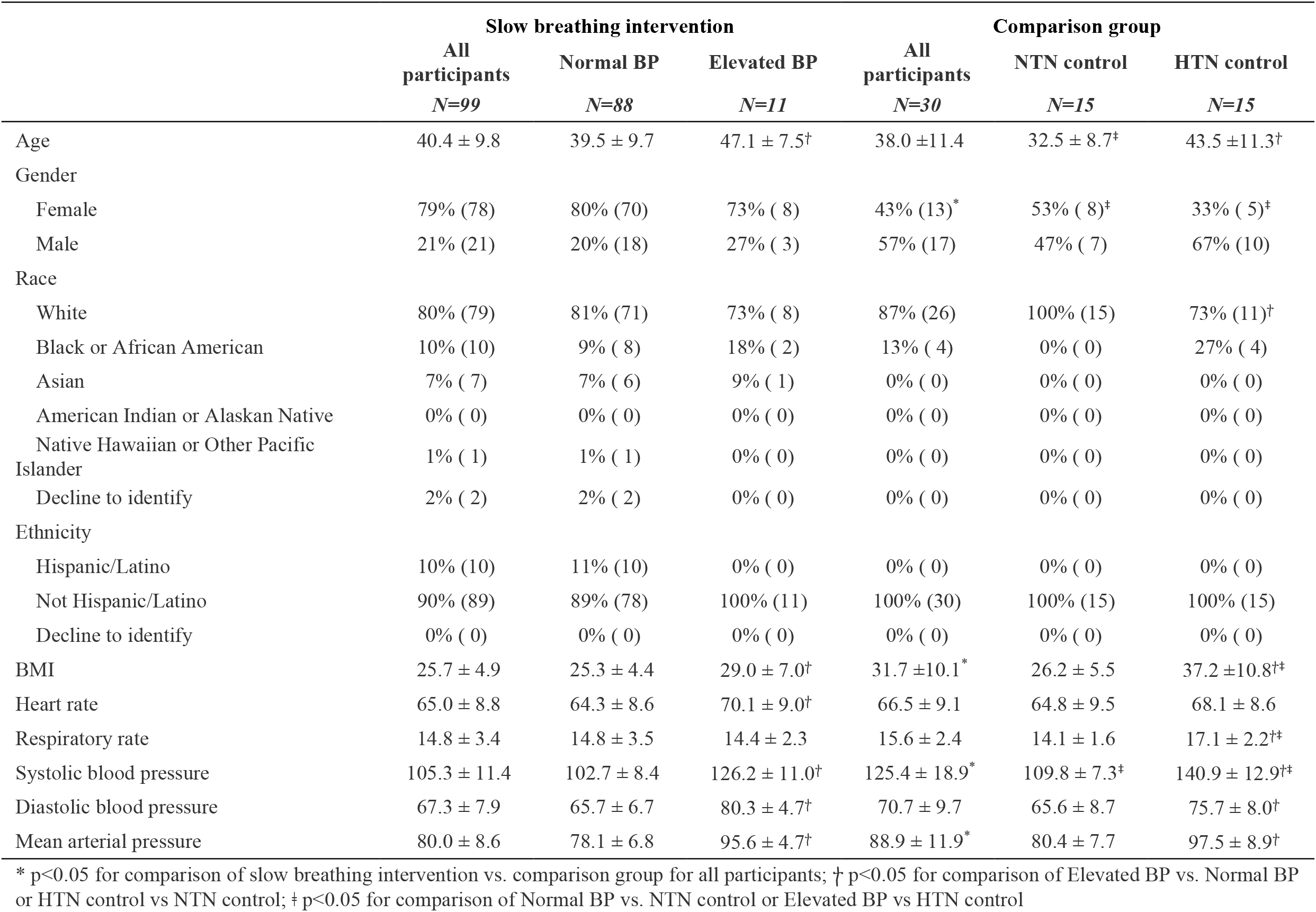

Changes in blood pressure are shown in Table 2 and Figure 2 for slow breathing study participants and the comparison group. SBP and DBP decreased significantly (−2.4 ± 7.3 mm Hg and -1.6 ± 5.5, respectively) at 12 weeks for all participants who practiced slow breathing. The comparison group showed no significant changes in blood pressure. Blood pressure reduced further among slow breathing participants with baseline SBP over 120mm Hg and/or DBP over 90mm Hg (−10.3 ± 7.9 and -3.8 ±5.5 mm Hg, respectively). In our regression model, higher SBP led to greater reduction in SBP from baseline to 12 weeks, (Figure 3). In the regression mode for DBP, baseline DBP was not associated with changes in DBP. There were no significant differences in blood pressure changes by breathing technique (E>I versus E=I) among all slow breathing participants or by sub-group analyses (normal blood pressure or elevated blood pressure).

**Table 2.**

|  | Slow breathing intervention |  |  | Comparison group |  |  |
| --- | --- | --- | --- | --- | --- | --- |
|  | All participants<br>*N=99* | Normal BP<br>*N=88* | Elevated BP<br>*N=11* | All participants<br>*N=30* | NTN control<br>*N=15* | HTN control<br>*N=15* |
| Systolic blood pressure, (mm Hg) |  |  |  |  |  |  |
| Baseline | 106.0 ± 11.5 | 103.2 ± 8.3 | 126.2 ± 11.0 | 125.4 ± 18.9 | 109.8 ± 7.3 | 140.9 ± 12.9 |
| Post | 103.6 ± 10.6 | 102.0 ± 9.8 | 115.9 ± 7.4 | 126.8 ± 20.9 | 109.4 ± 8.7 | 144.1 ± 13.6 |
| Change | -2.37 ± 7.34 | -1.29 ± 6.60 | -10.27 ± 7.86<sup>†</sup> | 1.36 ± 6.68<sup>*</sup> | -0.42 ± 4.18 | 3.13 ± 8.26<sup>‡</sup> |
| Diastolic blood pressure, (mm Hg) |  |  |  |  |  |  |
| Baseline | 67.6 ± 8.0 | 65.9 ± 6.7 | 80.3 ± 4.7 | 70.7 ± 9.7 | 65.6 ± 8.7 | 75.7 ± 8.0 |
| Post | 66.1 ± 7.8 | 64.6 ± 7.1 | 76.5 ± 4.0 | 68.7 ± 11.1 | 62.9 ± 10.3 | 74.5 ± 8.6 |
| Change | -1.57 ± 5.50 | -1.26 ± 5.47 | -3.82 ± 5.49<sup>†</sup> | -1.95 ± 7.76 | -2.71 ± 6.23 | -1.20 ± 9.20 |
| Mean Arterial Pressure, (mm Hg) |  |  |  |  |  |  |
| Baseline | 80.4 ± 8.7 | 78.3 ± 6.9 | 95.6 ± 4.7 | 88.9 ± 11.9 | 80.4 ± 7.7 | 97.5 ± 8.9 |
| Post | 78.6 ± 8.3 | 77.1 ± 7.5 | 89.6 ± 4.5 | 88.1 ± 13.4 | 78.4 ± 9.1 | 97.7 ± 9.4 |
| Change | -1.84 ± 5.13 | -1.27 ± 4.96 | -5.97 ± 4.63<sup>†</sup> | -0.85 ± 6.77 | -1.94 ± 4.98 | 0.24 ± 8.21 |

\* $p < 0.05$ for comparison of slow breathing intervention vs. comparison group for all participants; <sup>†</sup> $p < 0.05$ for comparison of Elevated BP vs. Normal BP or HTN control vs NTN control; <sup>‡</sup> $p < 0.05$ for comparison of Normal BP vs. NTN control or Elevated BP vs HTN control. Comparisons only made for pre-post changes.

**Figure 2.**
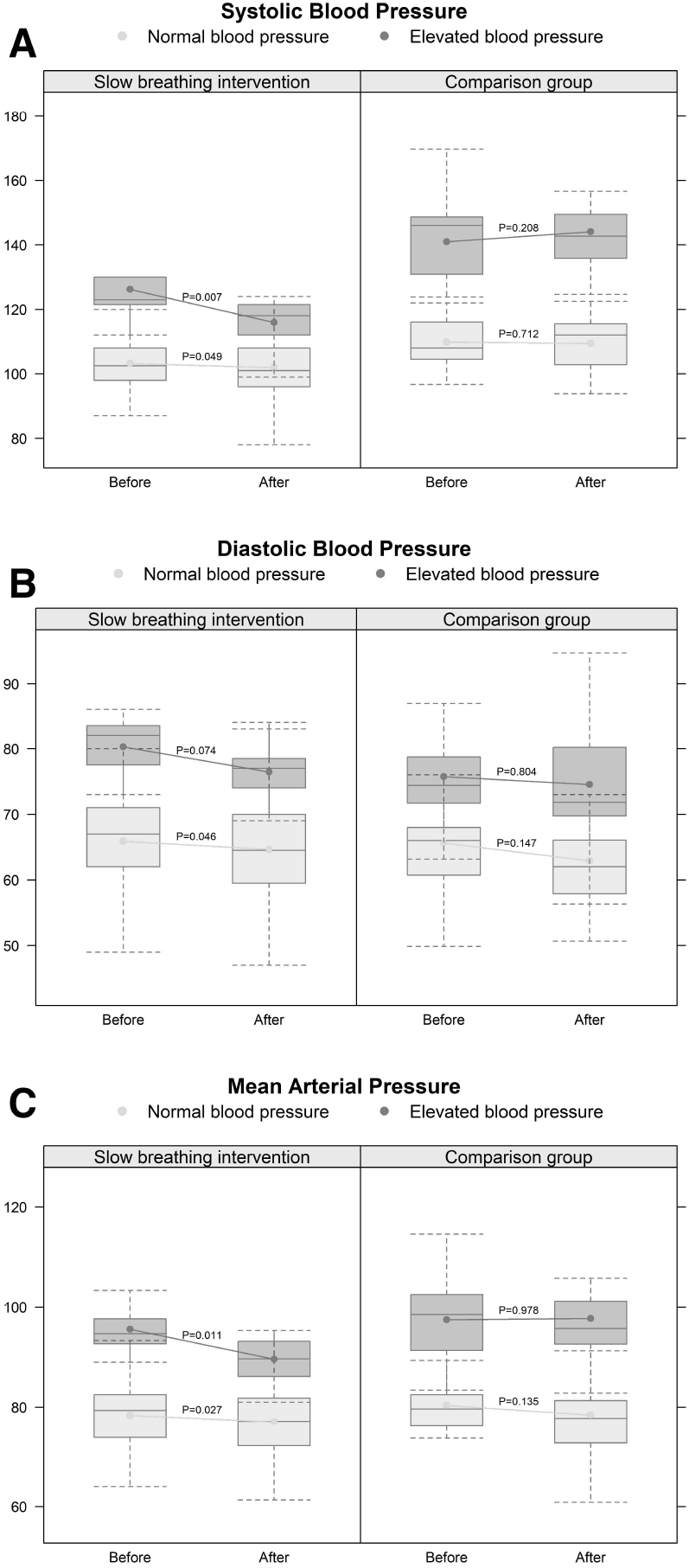
Twelve weeks of slow breathing training resulted in a significant decrease in systolic (Panel A), diastolic (Panel B) and mean (Panel C) arterial blood pressure among all subjects, but especially among subjects with elevated blood pressure (darker boxes), as compared with subjects with normal blood pressure (lighter boxes). For the comparison group, there were no differences observed for any of the parameters measured. p-values are based on Wilcoxon signed rank test.

**Figure 3.**
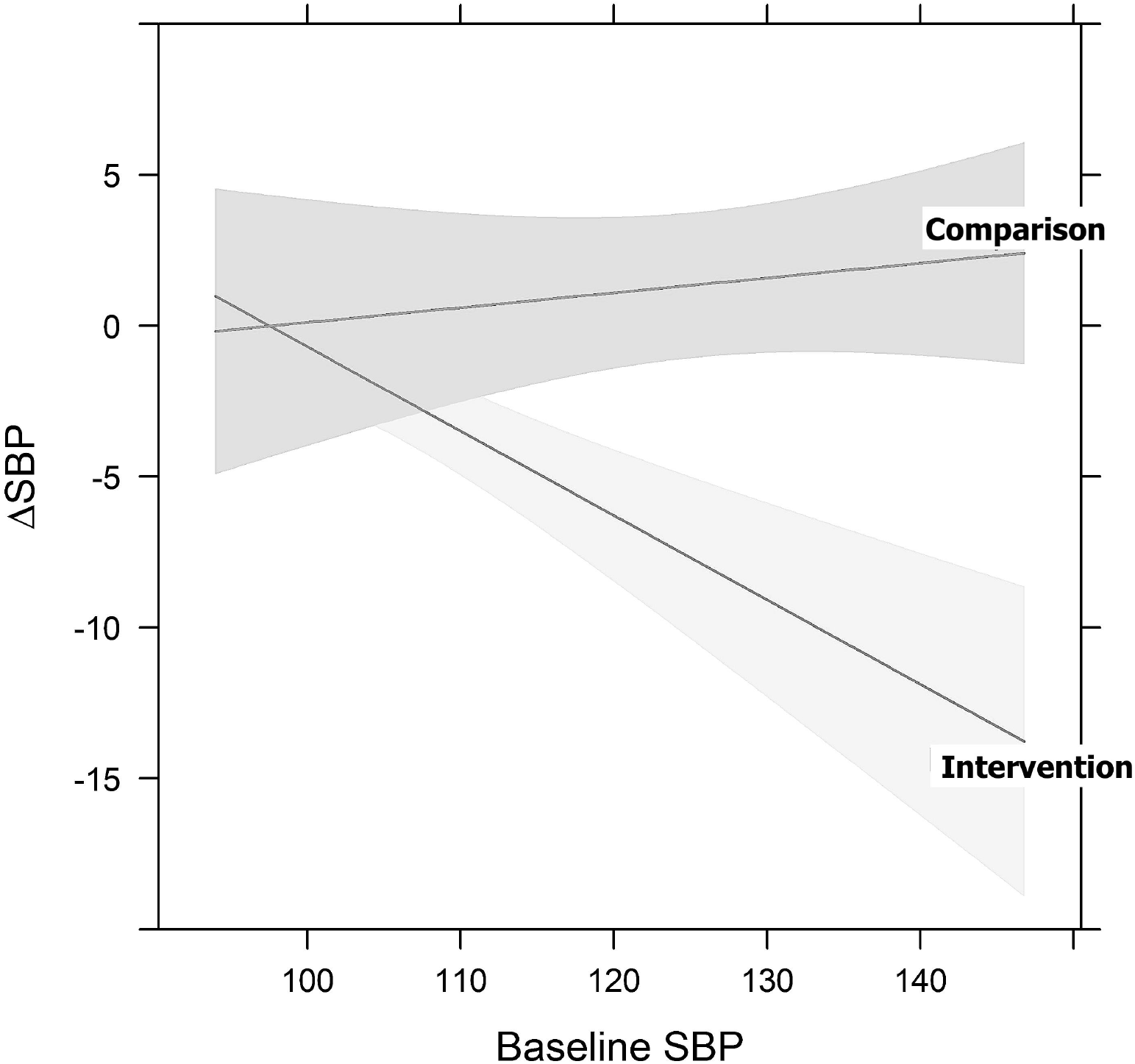
In the regression model of delta of SBP on baseline SBP, groups as well as interaction between baseline SBP and groups, we detected a significant association between baseline SBP and the predicted delta of SBP (based on the linear regression model) in the intervention group (darker shade), and no association in the comparison group (lighter shade).

Spectral analysis changes for slow breathing participants are shown in Table 3. We found that as expected at baseline, the high blood pressure groups (both intervention and comparison) had lower high frequency variability of heart rate, suggesting a relative impairment in cardiac parasympathetic modulation ^17–19^. The high blood pressure groups both in the intervention and control arms, also showed lower baroreflex sensitivity at baseline, also a confirmatory finding of dysregulation of the autonomic nervous system in hypertension ^20^. No significant differences were observed in spectral analysis changes from baseline to 12 weeks with the intervention. We found poor correlations between changes in blood pressure (SBP, DBP, MAP) and changes in autonomic measures including LF_SBP_ and HF_RR_.

**Table 3.**

|  | Slow breathing intervention |  |  | Comparison group |  |  |
| --- | --- | --- | --- | --- | --- | --- |
|  | All participants<br>N=99 | Normal BP<br>N=88 | Elevated BP<br>N=11 | All participants<br>N=30 | NTN control<br>N=15 | HTN control<br>N=15 |
| BRS HF (ms/mm Hg) |  |  |  |  |  |  |
| baseline | 20.7 ±17.9 | 22.1 ±18.5 | 10.7 ± 6.4 † | 18.4 ±18.5 | 28.1 ±21.2 | 8.7 ± 7.4 † |
| 12-week | 23.5 ±23.9 | 24.9 ±24.9 | 13.2 ±10.1 † | 16.8 ±17.1 * | 26.1 ±19.9 | 7.5 ± 5.0 † |
| change | 2.74 ±22.64 | 2.77 ±24.03 | 2.51 ± 7.08 | -1.58 ±11.12 | -1.99 ±14.01 | -1.16 ± 7.72 |
| BRS LF (ms/mm Hg) |  |  |  |  |  |  |
| baseline | 11.7 ± 7.9 | 12.2 ± 8.2 | 7.8 ± 3.8 † | 9.9 ± 6.6 | 12.5 ± 7.3 | 7.3 ± 4.6 † |
| 12-week | 12.7 ± 8.1 | 13.3 ± 8.3 | 7.9 ± 4.8 † | 10.0 ±10.7 * | 12.2 ±14.0 | 7.8 ± 5.3 |
| change | 0.983 ± 7.400 | 1.107 ± 7.801 | 0.082 ± 3.335 | 0.140 ± 9.879 | -0.288 ±12.198 | 0.568 ± 7.279 |
| SD<sub>RRI</sub> (ms<sup>2</sup>) |  |  |  |  |  |  |
| baseline | 55 ±31 | 58 ±32 | 38 ±12 † | 56 ±33 | 68 ±38 | 45 ±23 |
| 12-week | 61 ±34 | 63 ±36 | 48 ±17 | 64 ±68 | 85 ±91 | 44 ±22 |
| change | 6.07 ±24.90 | 5.54 ±26.16 | 9.88 ±12.45 | 8.04 ±58.42 | 16.93 ±73.73 | -0.86 ±38.27 |
| RMSSD (ms) |  |  |  |  |  |  |
| baseline | 47.8 ±41.1 | 51.0 ±42.8 | 25.1 ± 8.8 † | 47.1 ±40.8 | 60.5 ±48.4 | 33.6 ±26.9 † |
| 12-week | 51 ± 47 | 53 ± 49 | 33 ± 16 | 53 ± 78 | 78 ±106 | 28 ± 13 † |
| change | 2.959 ± 29.218 | 2.334 ± 30.832 | 7.501 ± 12.146 | 5.810 ± 57.726 | 17.056 ± 74.693 | -5.436 ± 32.443 |
| LF<sub>RRI</sub> (ms<sup>2</sup>) |  |  |  |  |  |  |
| baseline | 1034 ±1639 | 1103 ±1727 | 534 ± 540 | 1056 ±1326 | 1608 ±1686 | 504 ± 385 |
| 12-week | 1516 ±2035 | 1617 ±2129 | 782 ± 889 | 1624 ±4203 | 2684 ±5820 | 564 ± 562 |
| change | 483 ±1892 | 515 ±2009 | 247 ± 503 | 568 ±4103 | 1077 ±5809 | 60 ± 758 |
| HF<sub>RRI</sub> (ms<sup>2</sup>) |  |  |  |  |  |  |
| baseline | 744 ±1222 | 820 ±1285 | 190 ± 106 † | 801 ±1368 | 1229 ±1761 | 374 ± 618 † |
| 12-week | 896 ±1672 | 985 ±1763 | 244 ± 269 | 654 ±1117 | 1132 ±1438 | 175 ± 170 † |
| change | 151.5 ±1323.4 | 164.9 ±1409.6 | 54.5 ± 232.6 | -147.9 ± 692.9 | -97.0 ± 740.2 | -198.8 ± 664.2 |
| LF<sub>RRI</sub> / HF<sub>RRI</sub> |  |  |  |  |  |  |
| baseline | 2.50 ±2.47 | 2.43 ±2.49 | 3.00 ±2.38 | 3.09 ±3.66 | 2.06 ±1.94 | 4.11 ±4.66 |
| 12-week | 4.37 ±6.20 | 4.36 ±6.46 | 4.43 ±3.92 | 2.87 ±2.65 | 1.99 ±1.42 | 3.75 ±3.29 |
| change | 1.870 ± 6.559 | 1.931 ± 6.830 | 1.427 ± 4.287 | -0.217 ± 2.735 | -0.067 ± 2.029 | -0.366 ± 3.365 |
| LF<sub>SBP</sub> (mmHg<sup>2</sup>) |  |  |  |  |  |  |
| Baseline | 7.5 ± 5.9 | 7.2 ± 5.7 | 9.6 ± 7.1 | 10.7 ± 9.9 | 10.1 ±11.6 | 11.4 ± 8.2 |
| 12-week | 8.6 ± 6.2 | 8.3 ± 6.1 | 11.0 ± 6.4 | 12.2 ±10.4 | 14.4 ±11.0 | 10.1 ± 9.7 |
| Change | 1.124 ± 7.150 | 1.092 ± 7.264 | 1.354 ± 6.575 | 1.458 ±13.025 | 4.278 ±14.329 | -1.362 ±11.359 |
| HF<sub>SBP</sub> (mmHg<sup>2</sup>) |  |  |  |  |  |  |
| baseline | 1.88 ±2.28 | 1.74 ±2.22 | 2.91 ±2.55 | 3.20 ±3.88 | 1.36 ±0.89 | 5.03 ±4.82 † |
| 12-week | 1.56 ±1.50 | 1.46 ±1.38 | 2.29 ±2.10 | 3.06 ±2.93 * | 1.79 ±1.45 | 4.34 ±3.49 † |
| change | -0.317 ± 2.311 | -0.275 ± 2.414 | -0.623 ± 1.390 | -0.134 ± 3.413 | 0.423 ± 1.572 | -0.692 ± 4.581 |

Values represent Mean ± S.D for all participants in both the slow breathing intervention and the comparison controls. \* p<0.05 for comparison of slow breathing intervention vs. comparison group for all participants; † p<0.05 for comparison of Elevated BP vs. Normal BP or HTN control vs NTN control; ‡ p<0.05 for comparison of Normal BP vs. NTN control or Elevated BP vs HTN control. BRS, baroreflex slope; HF, high frequency; SD standard deviation; RMSSD, square root of mean squared successive differences, RRI, R-R interval; SBP, systolic blood pressure.

## Discussion

Many adults with hypertension have inadequate control of blood pressure. Additional treatment options, including non-pharmacologic, are necessary to provide additional options to patients. Slow breathing techniques are the most common mind-body practice in the United States, but evidence regarding long-term effects on blood pressure are lacking. The purpose of the study was to assess the effect of long-term slow breathing training on blood pressure and to explore its association with changes in autonomic function. We found that in healthy adults twelve weeks of slow breathing exercises resulted in a significant decrease in blood pressure measurements. The blood pressure decrease was more pronounced among participants with elevated baseline blood pressure (SBP>120 and/or DBP>90) exceeding the minimum clinically important difference for blood pressure change. Changes in blood pressure were independent to slow breathing technique (E>I versus E=I). Changes in spectral analyses (HF RR and LF sys) showed poor correlation to changes in blood pressure.

Slow breathing has been implemented in various ways including specific rates, use of device to provide biofeedback, and as part of other mind-body interventions such as mindfulness, t’ai chi, and meditation. Unfortunately, most of these studies had a short duration (four to eight weeks) and were done on hypertensive subjects already on medications. Many studies have measured the acute effects of slow breathing on blood pressure with long-term effects not being well documented in high quality studies. A systematic review and meta-analyses examined the clinical effectiveness and quality of research regarding the effects of slow breathing on blood pressure among hypertensives. (Chaddha *et al*., 2019). Chaddha found 15 device-guided breathing and two yoga-based slow breathing studies examining effects on blood pressure. Nine studies lacked an active control group. The slow breathing rate varied between studies. Six studies were sponsored by the manufacturer of RESPeRate. Compliance to breathing interventions was very high among all trials with no adverse effects. The total estimated effect of slow breathing on SBP was -5.88 mm Hg [-8.03—3.73] with high heterogeneity among studies (heterogeneity index, I^2^=83%). Device-based slow breathing reduced blood pressure less than yoga-based (−5.28 versus -8.45 mm Hg). Device-based slow breathing reduced DBP by -2.97 mm Hg [-4.28—1.66].

Our study intervention varies as compared to prior studies in several ways. First, we screened patients to determine if they would be able to adhere and perform slow breathing. Second, slow breathing was titrated to the slowest comfortable breath rather than a fixed goal of breathing. This is consistent with how slow breathing is taught amongst the tradition of slow breathing in yoga (pranayama). Among participants with the highest baseline blood pressure in our study (SBP>120, or DBP>90), the reduction in blood pressure (SBP -10.3 ± 7.9, DBP -3.8 ±5.5 mm Hg was greater than estimated total effects reported by Chaddha et al. Our results have several limitations. The effects of slow breathing on blood pressure were not the primary outcome of our study. Also, we excluded patients with hypertension which may have reduced our power to see potential effects on blood pressure. Lastly, even though we did not include a normal breathing group to control to account for a possible placebo effect, we were able to compare changes in blood pressure to participants in another of our studies with a similar time design.

Our results confirm previous studies that show differences in autonomic modulation between groups with normal blood pressure and elevated blood pressure ^21^. We found that at baseline, high blood pressure groups have lower vagal tone (HF_RRI_). We also found differences in baroreflex sensitivity between groups. We did not, however, find any changes in autonomic modulation with slow breathing training. This may be due to the high variability of the measurement and low sample size.

Prior studies that have found an acute reduction in blood pressure with slow breathing have hypothesized that slow breathing reduces blood pressure through decreases in sympathetic activity and/or increases in vagal tone ^22–24^. In our results, spectral analyses did not indicate significant changes or correlation with blood pressure reduction. One possibility for these results is that spectral analyses do not capture changes in autonomic tone sufficiently for the slow breathing intervention, or that the small number of subjects with elevated blood pressure prevented us from seeing more dramatic changes that could occur in real hypertensive subjects. More definitive measurements of autonomic function including norepinephrine spillover or direct measurement of muscle sympathetic nerve activity may provide additional information. Another possibility is slow breathing reduces blood pressure by other mechanisms such as endothelial function, oxidative stress, renal mechanisms, or salt metabolism ^25^. Further research into how slow breathing reduces blood pressure is necessary.

The large reductions in blood pressure we observed support the need for a larger randomized controlled efficacy trial to further examine slow breathing interventions among hypertensive adults. There is a predominance of device-guided studies, and our study adds to the limited data available regarding effects of yoga-based slow breathing on blood pressure. This study is of public health importance given that small reductions in blood pressure are associated with significant improvement in health outcomes. In pharmacological studies, a 5mm Hg reduction in SBP reduces risk of major cardiovascular events by 10%^26^. Yet, anti-hypertensive medications have low adherence and side effects. Whereas the literature suggests high compliance to slow breathing intervention with no adverse effects. Lastly, the absence of changes in autonomic tone as measured by spectral analyses suggest that the cardiovascular mechanisms of slow breathing have yet to be well characterized. Further research into the mechanisms and effectiveness of slow breathing on cardiovascular health is needed.

## Data Availability

All data produced in the present study are available upon reasonable request to the authors

## Novelty and Significance

### What is New?

Our study showed that regular slow breathing exercises significantly reduced blood pressure in the absence of changes in autonomic tone measured by spectral analyses.

We confirm that increases in blood pressure are associated with changes in autonomic tone.

### What is Relevant?

There is a dire need to find new and safe (no side effects) ways to treat hypertension. Slow breathing may be an additional therapeutic option for adults with hypertension.

### Summary

Regular slow breathing practice for 12 weeks reduces blood pressure. There were no significant changes in spectral analyses for heart rate variability or significant correlations between spectral analyses and blood pressure changes.

